# Artificial Intelligence Sepsis Prediction Algorithm Learns to Say “I don’t know”

**DOI:** 10.1101/2021.05.06.21256764

**Authors:** Supreeth P. Shashikumar, Gabriel Wardi, Atul Malhotra, Shamim Nemati

## Abstract

Sepsis is a leading cause of morbidity and mortality worldwide. Early identification of sepsis is important as it allows timely administration of potentially life-saving resuscitation and antimicrobial therapy. We present COMPOSER (COnformal Multidimensional Prediction Of SEpsis Risk), a deep learning model for the early prediction of sepsis, specifically designed to reduce false alarms by detecting unfamiliar patients/situations arising from erroneous data, missingness, distributional shift and data drifts. COMPOSER flags these unfamiliar cases as ‘indeterminate’ rather than making spurious predictions. Six patient cohorts (515,720 patients) curated from two healthcare systems in the United States across intensive care units (ICU) and emergency departments (ED) were used to train and externally and temporally validate this model. In a sequential prediction setting, COMPOSER achieved a consistently high area under the curve (AUC) (ICU: 0.925–0.953; ED: 0.938–0.945). Out of over 6 million prediction windows roughly 20% and 8% were identified as ‘indeterminate’ amongst non-septic and septic patients, respectively. COMPOSER provided early warning within a clinically actionable timeframe (ICU: 12.2 [3.2 22.8] and ED: 2.1 [0.8 4.5] hours prior to first antibiotics order) across all six cohorts, thus allowing for identification and prioritization of patients at high risk for sepsis.

---

Sepsis is a dysregulated host response to infection causing life-threatening organ dysfunction^1^. Approximately one in three hospital deaths are attributable to sepsis^2^. While effective protocols exist for treating sepsis^3^, challenges remain in early and reliable detection of this condition^4^. In recent years, the increased adoption of electronic medical records (EHRs) in hospitals has motivated the development of machine learning-based surveillance tools for detection^5–9^ and prediction^10–15^ of sepsis. However, most existing published sepsis prediction models are either based on data from a single hospital^5,7–10,13,15^ or multiple hospitals from the same healthcare system^11,14^ where the care processes are mostly standardized. Three major barriers to the widespread adoption of these systems are 1) their lack of generalizability across institutions, 2) high false alarm rates which can lead to alarm fatigue, and 3) the risk of *automation bias*, wherein users tend to over-rely on the system output ‘as a heuristic replacement of vigilant information seeking and processing’^16,17^, which could lead to misdetection and potential patient harm. One of the main factors contributing to an algorithm’s false alarm rate is the shift in distribution of population (encountering unfamiliar patients). A recent study demonstrated that detecting outlier cases and showing users an outlier focused message better enabled them to detect and correct for potential spurious predictions by an AI model^18^.

In this work, we propose COMPOSER (Conformal Multidimension Prediction of Sepsis Risk), a novel deep learning model for predicting sepsis. COMPOSER achieves improved generalizability and low false alarm rates through a prediction scheme that statistically determines conformity with a predefined collection of representations (aka *trust set*), as a means to establish the ‘conditions for use’ of the algorithm under unseen prediction scenarios. The proposed COMPOSER model consists of three modules. The *first module* makes use of clinical variables and timing information about measurements to generate lower dimensional representations that are robust to patterns of data missingness and institution-specific workflow practices^19,20^. The *second module* includes a *conformal prediction*^21–24^ network, which provides a statistical framework for detecting out-of-distribution (i.e., *indeterminate*) samples during the risk assessment phase in a deployment environment. Two bags of data representations (aka, *trust sets*) are used to quantify explicitly the conformity of new patient-level feature vectors to the previously seen examples of septic and non-septic feature vectors within the development cohort. The conformal prediction allows the model to detect outlier inputs that do not satisfy the conditions for use of the algorithm, which are subsequently assigned to an indeterminate predicted label class. Figure S3 of Supplementary Material provides an illustration of scenarios under which a test sample is accepted or rejected by the conformal prediction module. The *third module* includes a sepsis predictor that is a feedforward neural network followed by a logistic regression. Figure 1 (Panel B) provides the overall schematic diagram of COMPOSER during the evaluation phase. More details about the development of COMPOSER can be found in the Methods section. We evaluate the performance of this model on six patient cohorts, using a modified Area Under the Curve (AUC) metric evaluated under a clinically relevant protocol, as described by Hyland et al.^25^ Additionally, we report threshold-based performance metrics (at a fixed sensitivity of 80%), with a focus on false alarms and misdetections, using positive predictive value (PPV) and negative predictive value (NPV), respectively.

**Figure 1:**
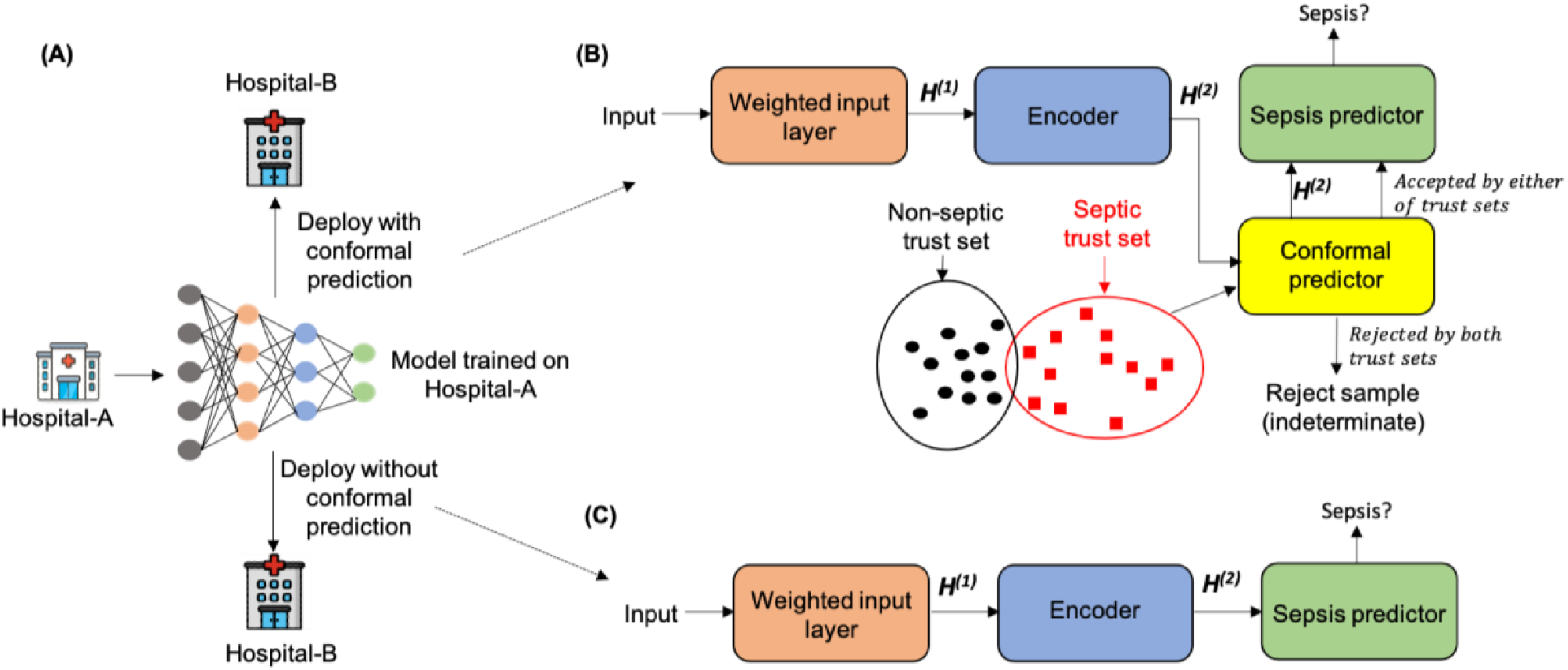
Schematic diagram of COMPOSER. During evaluation phase (Panel B), the input test data is first fed into the weighted input layer, and is then passed through the encoder (*first module*), afterwhich the method of conformal prediction is utilized to determine whether a trustworthy prediction can be made (*second module*). This is achieved by comparing the conformity of the new *de-nuanced* representation (*H*^(2)^) to the representations in the trust set, which are carefully selected during the training phase (see Methods section). If conformity can be achieved at a given confidence level (*ϵ*), *H*^(2)^ is then forwarded to the sepsis predictor to obtain a risk score (*third module*). In comparison, Panel C shows a deployment scheme without the use of conformal prediction.

COMPOSER was trained and evaluated on six patient cohorts (515,720 patients) across two academic medical centers (Hospital A, Hospital B) in the US, including three ICU cohorts and three ED cohorts collected between 2016 and 2020. The patient characteristics of all the six cohorts have been tabulated in Appendix B - Tables S2 and S3 of Supplementary Material. Patients in the Hospital-A ICU and ED cohorts were randomized across training (80%) and testing (20%) sets. The entire Hospital-A Temporal ICU and ED, and Hospital-B ICU and ED cohorts were used for temporal and external validation respectively. Patients 18 years or older were followed throughout their stay (ICU or ED depending on the cohort) until discharge or development of sepsis according to the latest International Consensus Definitions for Sepsis (Sepsis-3)^1,26^.

## Internal testing set

COMPOSER’s performance (AUC/PPV) on the source cohort ICU and ED testing dataset was 0.953/38% and 0.945/20.1%, respectively. COMPOSER was able to achieve this performance while maintaining low false alarms (false alarms per person hour of 0.031 and 0.042; SPC of 93.0% and 93.5%). Figures S7 and S8 of Supplementary Material show heatmaps of the top 15 variables contributing to the increase in risk score up to 12 hours prior to onset of sepsis. It was observed that clinical variables related to sepsis (e.g., temperature, white blood count, heart rate) were identified as the top contributing factors.

## External validation

When applying COMPOSER to data from Hospital-B ICU patients, compared to a baseline feedforward neural network (FFNN) model, COMPOSER achieved roughly 6.8 times reduction in false alarms per person hour (0.043 versus 0.296; SPC of 90.7% vs 67.0%) while maintaining superior AUC and PPV (AUC of 0.925 vs 0.910, p<0.001; PPV of 24.3% vs 23.0%). Similarly, within the Hospital-B ED cohort COMPOSER achieved roughly 4.5 times reduction in false alarms per person hour (0.038 versus 0.172 ; SPC of 94.7% vs 82.1%) while maintaining superior AUC and PPV (AUC of 0.938 vs 0.910, p<0.001; PPV of 13.4% vs 13.0%). See Figure 2, Panels A-F for a comparison of COMPOSER’s performance (PPV, NPV, DOR, SPC, AUC and False alarms per person hour) vs the baseline FFNN model across all cohorts.

**Figure 2:**
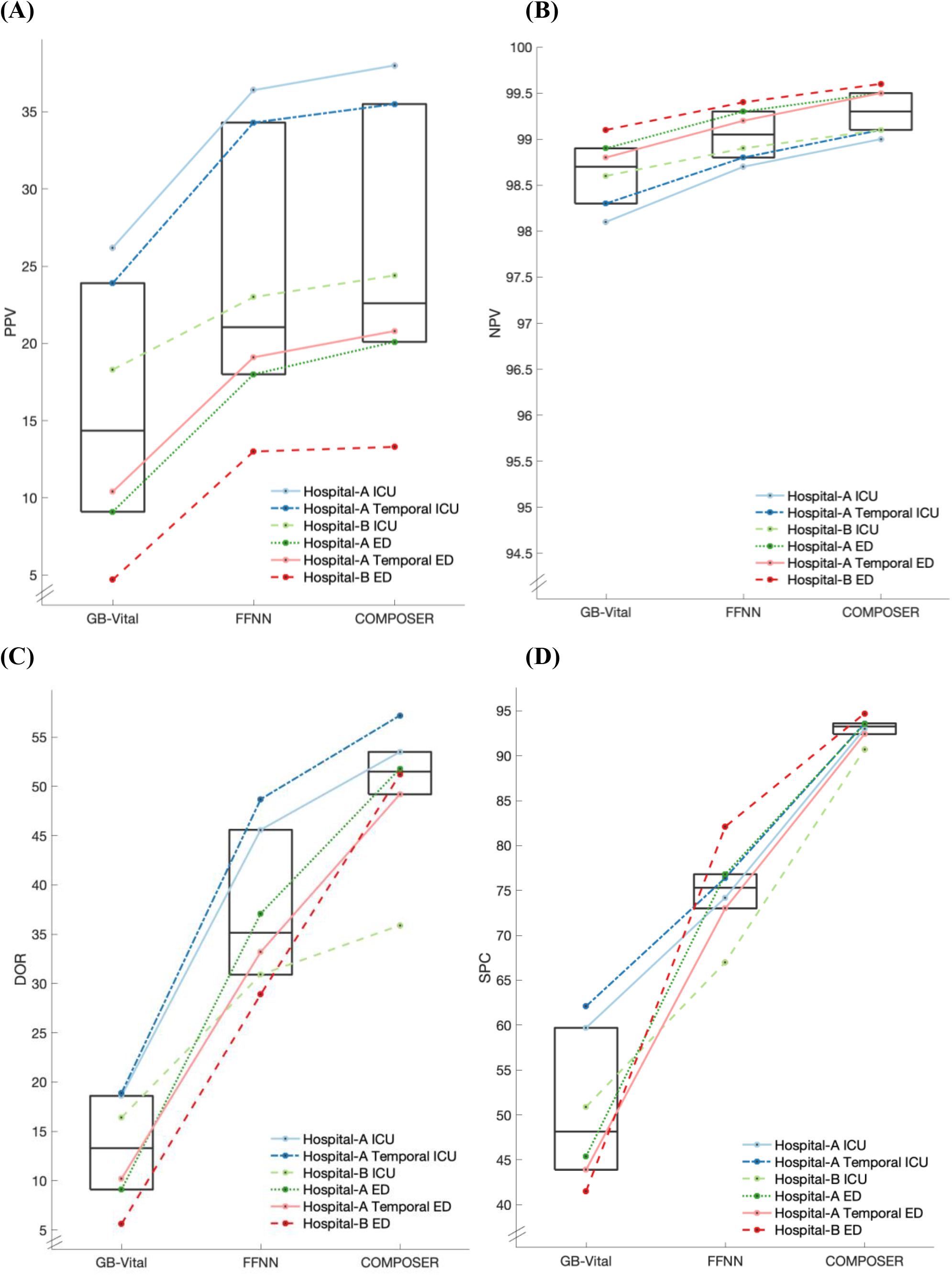
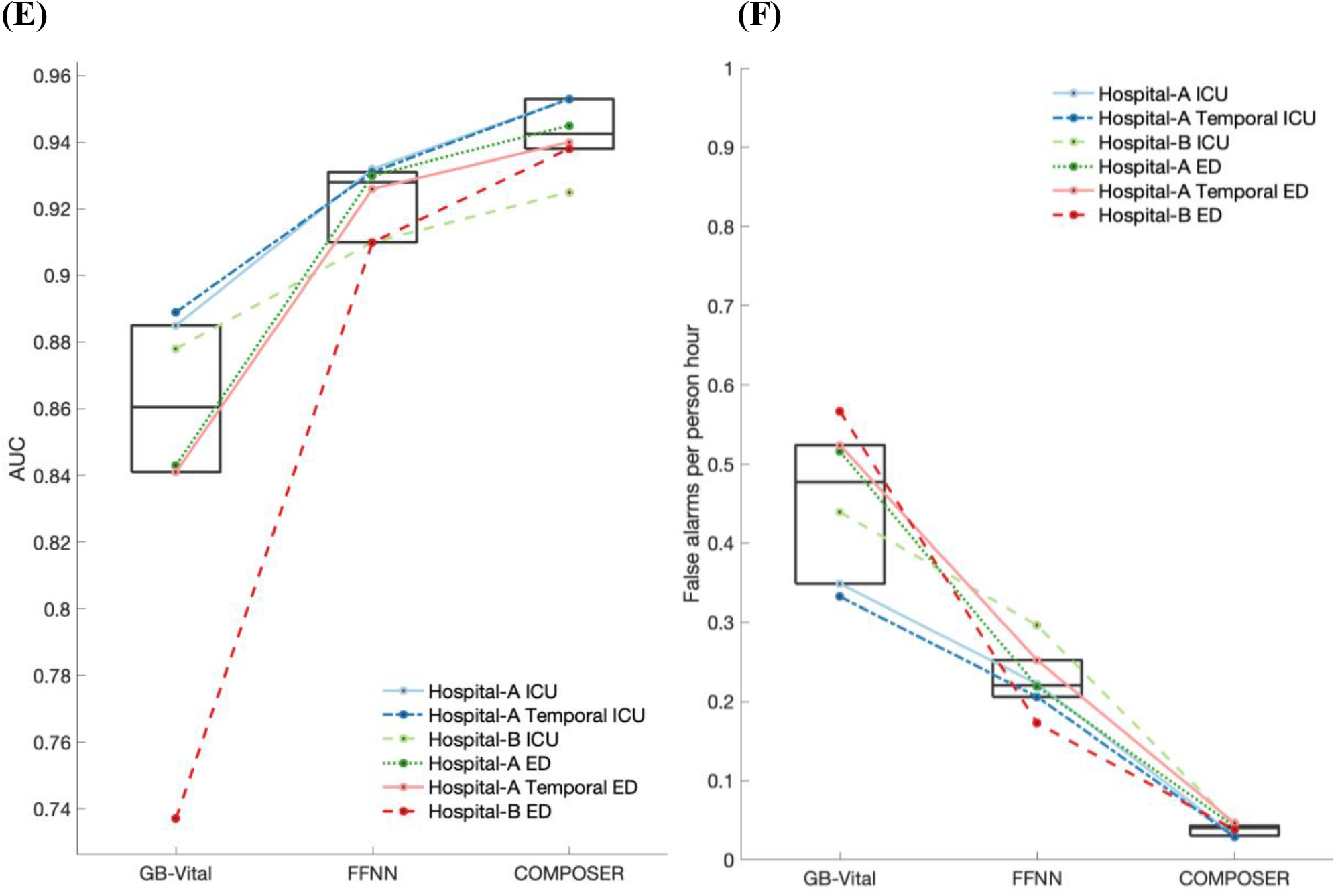
Summary of COMPOSER performance. Comparison of COMPOSER model against *GB-Vital*^a^ and a feedforward neural network (*FFNN*^b^). The line plots in Panels A-F show the relative improvement in positive predictive value (PPV), negative predictive value (NPV), diagnostic odds ratio (DOR), specificity (SPC), Area Under the Curve (AUC) and number of false alarms per person hour (FAPH), respectively. The median and interquartiles for all six cohorts (three ICUs and three EDs) are summarized via superimposed box plots. In comparison, *ESPM*^c^ (not shown here) achieved an AUC of 0.889 (PPV=31.2%, NPV=97.8%, DOR=23.2, SPC =84.3, FAPH=0.132) and 0.876 (PPV=35.9%, NPV=96.8%, DOR=17.4, SPC =94.2%, FAPH=0.05) across Hospital-A temporal ICU and ED. ^a^ *GB-Vital* corresponds to a Gradient Boosted Tree (XGBoost)^6,15^ built using six vital signs measurements: systolic blood pressure, diastolic blood pressure, heart rate, respiratory rate, oxygen saturation and temperature. ^b^ *FFNN* corresponds to a 2 layer feedforward neural network that uses the same number of input features as that of COMPOSER. The starting point of y-axis for Panels A and B were determined by the chance level of a classifier at the lowest prevalence rate. ^c^ ESPM corresponds to the Epic’s commercially available Best Practice Advisory (BPA) alert. We only had access to the risk scores produced by this system at Hospital-A during the temporal validation time-frame.

## Outlier Detection and Indeterminates

Overall 75-86% of the prediction windows (see Table S6 and Table S7 of Supplementary Material) satisfied the conditions for use of the algorithm, and therefore the corresponding predictions were deemed trustworthy. However, within the septic windows the percentage of indeterminate cases were almost half compared to the non-septic windows (24.6%-27.8% versus 12.4%-13.9% for the ICU cohorts and 13.6%-16.1% versus 6.5%-9.9% for the ED cohorts). Moreover, patient-wise analysis revealed that the median percentage of septic patients with all septic windows rejected was 1.1% [0.87%-2.6% IQR], indicating that conformal prediction had minimal deleterious effect on the patient-wise sensitivity of the algorithm.

## Temporal validation

To assess the impact of changes in institutional practices and patient populations over time we performed an experiment in which a model trained on a combined cohort of ICU and ED patients (which consisted of patients admitted from January 2016 through March 2019) from Hospital-A was applied to a temporal validation set consisting of patients admitted to Hospital-A from August 2019 through February 2020. COMPOSER achieved an AUC of 0.940 (PPV of 20.8%) on the Hospital-A Temporal ED validation cohort and an AUC of 0.952 (PPV of 35.5%) on the Hospital-A Temporal ICU validation cohort (see Table 2). COMPOSER was able to achieve this performance while maintaining low false alarms; false alarms per person hour of 0.047 (SPC of 92.4%) and 0.029 (SPC of 93.6%) on the temporal ED and ICU cohorts, respectively. This finding suggested that the model was not adversely impacted by changes in patient population or clinical practices over time. In comparison, a commercially available sepsis prediction model (ESPM) in use at Hospital-A during the same time period had roughly 4.5 times higher false alarms per person hour in the ICU (0.132 vs 0.029). Moreover, COMPOSER achieved significantly better alarm notification lead-time, in advance of clinical suspicion of sepsis, compared to the ESPM model (see Table 1).

**Table 1:** Time from model prediction to clinical suspicion of infection. The difference between the time at which an alarm (*t*^*alarm*^) was fired (i.e. risk score exceeded a decision threshold) and *t*_*lactate*_ */t*_*suspicion*_*/t*_*ABX*_*/t*_*culture*_ for the Hospital-A Temporal ICU and ED cohorts are shown. *t*_*alarm*_ corresponded to the earliest time at which an alarm was fired in the interval [*t*_*suspicion*_-12 hours, *t*_*suspicion*_+ 12 hours]. *t*_*lactate*_ corresponded to the earliest time at which a lactate measurement was made in the interval [*t*_*suspicion*_-12 hours, *t*_*suspicion*_+ 12 hours]. *t*_*ABX*_ corresponded to the time at which antibiotics were ordered following *t*_*suspicion*_. *t*_*culture*_ corresponded to the time at which cultures were ordered following *t*_*suspicion*_. A negative value indicates the alarm was fired after the time point of interest.

| | $\Delta t_{lactate}$<br>(in hours) | $\Delta t_{suspicion}$<br>(in hours) | $\Delta t_{ABX}$<br>(in hours) | $\Delta t_{culture}$<br>(in hours) |
| --- | --- | --- | --- | --- |
| **Hospital-A Temporal ICU** |  |  |  |  |
| **COMPOSER** | 2 [0 7] | 8.4 [1.2 11.5] | 12.2 [3.2 22.8] | 11.2 [5.2 11.9] |
| **ESPM (score>5)** | 1 [0 5] | 3.5 [0.2 11.2] | 10.2 [1.0 18.9] | 8.9 [1.2 11.9] |
| **Hospital-A Temporal ED** |  |  |  |  |
| **COMPOSER** | 1 [1 2] | 1.0 [0.3 2.3] | 2.1 [0.8 4.5] | 0.8 [0.2 3.4] |
| **ESPM (score>5)** | 0 [-1 1] | 0.1 [-0.9 0.65] | 0.6 [0.2 2.0] | 0.2 [-0.8 1.0] |

**Table 2:** Summary of performance of COMPOSER on the (A) ICU and (B) ED cohorts when evaluated in a sequential prediction setting. Decision threshold corresponding to 80% sensitivity on the Hospital-A training cohort (0.561 and 0.414 for the ICU and ED cohorts respectively).

**(A) ICU cohorts**
|  | SEN | SPC | PPV | NPV | DOR | AUC |
| --- | --- | --- | --- | --- | --- | --- |
| **Hospital-A ICU test**<br>N<sup>*</sup>=3,406; S<sup>*</sup>=767 (22.5%) | 91.6<sup>*</sup> | 93.0<sup>**</sup> | 38.0<sup>**</sup> | 98.8<sup>**</sup> | 53.5<sup>**</sup><br>(1.088) | 0.953 |
| N<sup>**</sup>=157,527; S<sup>**</sup>=6499 (**4.1%**) | 80.1<sup>**</sup> |  |  |  |  |  |
| **Hospital-A Temporal ICU**<br>N<sup>*</sup>=3,596; S<sup>*</sup>=733 (20.4%) | 92.3<sup>*</sup> | 93.6<sup>**</sup> | 35.5<sup>**</sup> | 99.0<sup>**</sup> | 57.2<sup>**</sup><br>(1.090) | 0.953 |
| N<sup>**</sup>=171,242; S<sup>**</sup>=6156 (**3.6%**) | 79.3<sup>**</sup> |  |  |  |  |  |
| **Hospital-B ICU**<br>N<sup>*</sup>=45,812; S<sup>*</sup>=7,913 (17.3%) | 91.3<sup>*</sup> | 90.7<sup>**</sup> | 24.3<sup>**</sup> | 99.1<sup>**</sup> | 35.9<sup>**</sup><br>(1.027) | 0.925 |
| N<sup>**</sup>=1,884,383; S<sup>**</sup>=56,637(**2.9%**) | 78.9<sup>**</sup> |  |  |  |  |  |

**(B) ED cohorts**
|  | SEN | SPC | PPV | NPV | DOR | AUC |
| --- | --- | --- | --- | --- | --- | --- |
| **Hospital-A ED test**<br>N<sup>*</sup>=19,807; S<sup>*</sup>=1,624 (8.2%) | 95.6<sup>*</sup> | 93.5<sup>**</sup> | 20.1<sup>**</sup> | 99.5<sup>**</sup> | 51.8<sup>**</sup><br>(1.109) | 0.945 |
| N<sup>**</sup>=162,504 S<sup>**</sup>=7691 (**4.7%**) | 78.1<sup>**</sup> |  |  |  |  |  |
| **Hospital-A Temporal ED**<br>N<sup>*</sup>=19,945; S<sup>*</sup>=1,795 (9.0%) | 96.0<sup>*</sup> | 92.4<sup>**</sup> | 20.8<sup>**</sup> | 99.5<sup>**</sup> | 49.2<sup>**</sup><br>(1.107) | 0.940 |
| N<sup>**</sup>=158,045; S<sup>**</sup>=8766 (**5.5%**) | 80.0<sup>**</sup> |  |  |  |  |  |
| **Hospital-B ED**<br>N<sup>*</sup>=330,299; S<sup>*</sup>=14,454 (4.4%) | 90.5<sup>*</sup> | 94.7<sup>**</sup> | 13.4<sup>**</sup> | 99.6<sup>**</sup> | 51.2<sup>**</sup><br>(1.031) | 0.938 |
| N<sup>**</sup>=2,362,426; S<sup>**</sup>=64032(**2.7%**) | 70.5<sup>**</sup> |  |  |  |  |  |

N: Number of patients within the cohort; S: Number of septic patients
\*Patient-wise ; \*\* hourly window-wise
**SEN:** Sensitivity, **SPC:** Specificity, **PPV:** Positive predictive value, **NPV:** Negative predictive value, **DOR:** Diagnostic odds ratio with 95% confidence interval

## Analysis of missed-detections

We observed that COMPOSER achieved high negative predictive value across all the six cohorts (98.8%–99.1% in ICU and 99.5–99.6% in ED) when evaluated in a sequential prediction setting. Additionally, it was also observed that COMPOSER made at least one positive prediction within the septic window in 91.3%–92.3% and 90.5%–95.6% of septic patients in the ICU and ED cohorts respectively. In other words, COMPOSER’s missed-detection rate amongst septic patients was below 10% across all the six cohorts. Thus mitigating concerns about deleterious effects of automation bias; in particular, when used in association with the standard of care and existing hospital workflow practices for identification of sepsis.

In summary, we presented a generalizable deep learning model for the continuous prediction of sepsis within a clinically actionable window of 2-24 hours in advance of clinical recognition. Using representation learning and conformal prediction allowed us to introduce a formal procedure to detect outlier inputs (or unfamiliar patients/situations) arising from erroneous data, missingness, distributional shift and data drifts, and thus establishing the ‘conditions for use’ of the algorithm. By carefully constructing two trust-sets for the representation of septic and non-septic data points by, for example, varying tolerance for missing data (see Appendix E of Supplementary Material for construction of trust-sets), conformal prediction was able to control the rejection rate of septic and non-septic cases independently. As such, labeling outlier inputs as ‘indeterminates’ resulted in improvement in the rate of false alarms by a factor of 5-10 compared to baseline models, while keeping sepsis missed-detection rates among the indeterminate cases close to zero. In addition to the improved PPV, COMPOSER’s NPV remains high (98.8% ICU and 99.5% ED), thus mitigating concerns around potential missed-detections and automation bias when used in parallel with the standard of care for detection of sepsis. Recent literature demonstrates that flagging outlier cases in predictive tools is more likely to enable the end-users to detect and correct for potential model mistakes^18^. Flagging of indeterminate cases can be incorporated into a verification process^27^ to elucidate the underlying causes of indeterminacy and to take corrective actions (e.g., ordering of additional labs or fine-tuning of the model on the target population^28^). We show consistent performance across different levels of care, hospitals, and retrospective and temporal validation cohorts, demonstrating that our approach is broadly applicable to different care settings. Additionally, COMPOSER was designed to be locally interpretable wherein the model was capable of identifying the most relevant features contributing positively or negatively to the increase in risk score at every point in a patient’s timeline. To maximize predictive performance, input to such algorithms should be enriched with higher resolution data from bedside monitors and biomarkers for pathogen profiling and host inflammatory response to infection. The proposed algorithm provides a foundational building block for the design of future generalizable and trustworthy prediction tools potentially useful in other clinical syndromes and disease processes. A real-time HL7 FHIR^29,30^ compatible software pipeline has been developed to enable interoperable multi-center deployment of this algorithm (See Appendix H of Supplementary Material for more details). Future work includes performing prospective clinical trials to validate COMPOSER’s predictions in a real-time clinical setting. However, our findings provide significant clinical evidence for a radical improvement in early identification of sepsis, without incurring unduly cognitive burden on caregivers, and has the potential to improve sepsis-related clinical outcomes.

## Data Availability

De-identified data from the Emory cohort has been made available as part of the PhysioNet Challenge 2019. Access to de-identified UCSD cohort may be made available via approval from UCSD Institutional Review Board (IRB) and Health Data Oversight Committee (HDOC). Access to the computer code used in this research is available upon request to the corresponding author.

## Author Contributions

S.P.S and S.N were involved in the original conception and design of the work, developed the network architectures, conducted the experiments and analyzed the data. GW and AM provided clinical expertise, reviewed patient data and contributed to interpretation of results and the write-up. S.P.S prepared all the figures. SPS, GW AM and S.N wrote and edited the final manuscript.

## Methods

### Dataset description

We collected de-identified data from the EHR across different care-levels and time-frames from two academic medical centers (Hospitals A and B) in the United States to construct a total of six patient cohorts (total of 515,720 encounters). Appendix B of Supplementary Material contains detailed information regarding the six cohorts considered in this study. Patients 18 years or older were followed throughout their stay until development of sepsis or discharge. To allow for initial examination and stabilization of patients and adequate data collection for prediction purposes, we focused on sequential hourly prediction of sepsis starting at hours two and four within our ED and ICU cohorts, respectively. Patients who were identified as having sepsis prior to prediction start time or those with no measurement of heart rate or blood pressure prior to the prediction start time or those whose length of stay (ICU or ED depending on the cohort) was more than 21 days were excluded.

The overall dataset was divided into the following cohorts: 1) *training cohort* (80% of encounters from Hospital A ICUs and EDs, including over 13,000 ICU encounters and over 79,000 ED encounters), 2) *testing cohort* (20% of the encounters from Hospital A ICUs and EDs), 3) *temporal validation cohort* (prospectively collected encounters from Hospital A ICUs and EDs), and 4) *external validation cohort* (retrospective data from Hospital B ICUs and EDs). See Table 2 for a breakdown of patients used in cohorts 2-4 for evaluation purposes after the model was trained on cohort 1 and was frozen. The Hospital-A training and testing cohorts were collected between January 2016 and August 2019, and the temporal validation cohort was collected between August 2019 through February 2020. The external validation cohort was collected between January 2014 to December 2018.

We followed the latest guidelines provided by the Third International Consensus Definitions for Sepsis (Sepsis-3)^1,26^ which defined sepsis as a life-threatening organ dysfunction caused by a dysregulated host response to infection. As such, the two main criteria for establishing onset time of sepsis included: 1) evidence of acute organ dysfunction, and 2) evidence of infection. This investigation was conducted according to University of California San Diego IRB approved protocol #191098 and Emory University IRB Protocol #00110675.

### Model Development

Consistent with the PhysioNet Sepsis Challenge 2019, a total of 40 clinical variables (34 dynamic and 6 demographic variables, see Table S1 of Supplementary Material) were extracted based on their association with the onset of sepsis and their availability in EHR across the two hospitals considered in our study^11,12,14^. These included vital signs measurements (heart rate, pulse oximetry, temperature, systolic blood pressure, mean arterial pressure, diastolic blood pressure, respiration rate and end-tidal carbon dioxide), laboratory measurements (bicarbonate, bicarbonate excess, fraction of inspired oxygen, pH, partial pressure of carbon dioxide from arterial blood, oxygen saturation from arterial blood, asparate transaminase, blood urea nitrogen, alkaline phosphatase, calcium, chloride, creatinine, bilirubin direct, serum glucose, lactate, magnesium, phosphate, potassium, total bilirubin, troponin, hematocrit, hemoglobin, partial thromboplastin time, leukocyte count, fibrinogen and platelets) and demographic variables (age, gender, identifier for medical ICU unit, identifier for surgical ICU unit, length of hospital stay, length of ICU stay). All vital signs and laboratory variables were organized into 1-hour non-overlapping time series bins to accommodate for different sampling frequencies of available data. All the variables with sampling frequencies higher than once every hour were uniformly resampled into 1-hour time bins, by taking the median values if multiple measurements were available. Variables were updated hourly when new data became available; otherwise, the old values were kept (sample-and-hold interpolation). Mean imputation was used to replace all remaining missing values (mainly at the start of each record). Additionally, for every vital signs and laboratory variable, their local trends (slope of change), baseline value (mean value measured over the previous 72 hours) and the time since the variable was last measured (TSLM) were recorded. Hereafter, we refer to the 34 dynamical variables and their local trends (total of 102 features) by *X*_*dyamical*_, the 34 TSLM features by *X*_*TSLM*_ and the 6 covariate features by *X*_*covar*_, resulting in a total of 108 features.

### Development of the COMPOSER model

COMPOSER consists of three modules. First, a *weighted input layer* that scales the value of a clinical variable depending on the time since it was last measured. Intuitively, this layer attempts to mimic a clinician’s thought process of putting more importance on the most updated vitals and labs, depending on the physiologically plausible rates at which such measurements can change. As such, the weighted input layer was designed to incorporate some information about the timing of clinical measurements without allowing the network to exploit the correlation between frequency of measurements and disease severity or patients’ level of care^31^. Such factors are often affected by the institution-specific workflow practices and care protocols and likely to reduce the generalizability of a predictive algorithm. The output of this layer was fed into an *encoder network* (a feed forward neural network) that is used to reduce data dimensionality. The second module is a *conformal predictor* which is used to establish the ‘conditions for use’ of the model by statistically assessing the conformity of any new test instance to a pre-constructed bag of examples (‘trust set’) drawn from the training set. The third module includes a sepsis predictor, which is a feedforward neural network whose output is a probability score (between 0 and 1) that represents the risk of onset of sepsis within the next four hours. All the modules are parametrized as neural networks and trained end-to-end, and enable the application of local interpretability methods such as relevance scores (RS) and layer-wise relevance propagation (LRP)^32^.

Conformal prediction enables the algorithm to determine the level of data covariance shift at which one may still trust a clinical risk score. The development and evaluation of COMPOSER involved two steps: First, the first and the second modules were trained using the combined Hospital-A ICU and ED training sets. Second, the trust set (consisting of representations from the encoder module) was constructed from the combined Hospital-A ICU and ED training sets. Finally, the trained modules along with the trust set were used for model evaluation. Each of the individual components of COMPOSER, namely the weighted input layer and conformal prediction are explained in detail in the following sections.

### Weighted input layer

We designed a weighted input layer that scales the latest measured value of a variable depending on the duration since it was measured. This scaling enables the model to appropriately account for the age of an imputed feature while constraining the model from directly exploiting the frequency of measurements. The extent of scaling is controlled by a parameter *α* that is learned from data. Let us consider 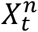 to be a centered and standardized 68 dimensional vector (consisting of all dynamical variables) at time *t* for patient *n*. Henceforth, with a slight abuse of notation we will refer to 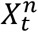 by *X*, wherein *X* = [x_1_; x_2_; ….; x_68_], x_j_ ∈ ℝ. Next *δ*_*j*_ corresponds to the duration since variable *j* was last measured (in reference to the current time *t*). Each of the variables *x*_*j*_ is then non-linearly weighted based on the duration since it was last measured, to obtain 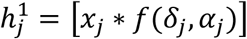. The weighting function *f*(.) is defined as follows:

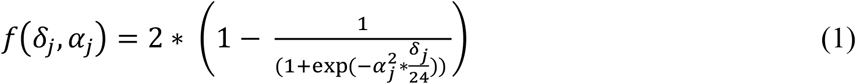

Where *α*_*j*_. is a scaling factor for each of the dynamical variables and is learned during the training of the model. Thus, the output of the weighted input layer is a 68 dimensional feature vector 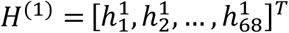. The scaling factors *α*_*j*_ obtained at the end of model training is inversely related to the extent of sample-and-hold interpolation that is useful for the i-th variable.

### Detecting distribution shift using conformal prediction

We used the method of *conformal prediction*^21,23,24^ to develop a statistical test to determine whether a given data sample belongs to the data distribution from which the training data was drawn. A sepsis prediction is made on the data sample only if it belongs to the training distribution of COMPOSER, else the data sample is rejected and no sepsis prediction is made. Two sets of size *M* of the de-nuanced representations 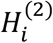 from the source dataset (*S*) each containing only septic and non-septic examples, respectively, are chosen as the trust sets 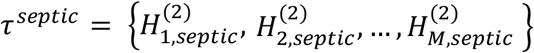 and 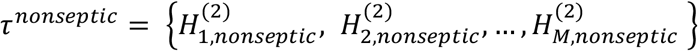. We are then given a test example 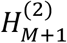 for which the task of conformal prediction is 1) to predict if the test example is drawn from the same probability distribution as that of other examples in the either of the trust sets *τ*^*septic*^ and *τ*^*nonseptic*^ at a given confidence level (1 - *ε*), and 2) if yes, the test example 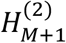 is passed onto sepsis predictor to obtain the sepsis risk score. We choose ε to be 0.05 in our analysis, which translates to 95% confidence that the new test example 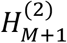 is drawn from the same distribution as that of the trust set *τ*^*septic*^ or *τ*^*nonseptic*^. See Appendix E of Supplementary Material for more details on the theory behind conformal prediction and construction of the trust sets.

### Statistical Methods and Hypothesis Testing

For all continuous variables, we have reported median ([25th – 75th percentile]). For binary variables, we have reported percentages. The area under receiver operating characteristic (AUC) curves statistics, specificity (SPC), positive predictive value (PPV), negative predictive value (NPV) and diagnostic odds ratio (DOR) at a fixed threshold (corresponding to 80% sensitivity level on the development cohort) were calculated to measure the performance of the models. Additionally, we report false alarms per person hour (FAPH) which can be used to calculate the expected number of false alarms per unit of time in a typical care unit (e.g., a FAPH of 0.025 translates to roughly 1 alarm every 2 hours in a 20-bed care unit). Statistical comparison of all AUC curves was performed using the method of DeLong et al.^33^ Statistical comparison of DOR was performed using the paired t-test. Additionally, as a secondary end-point we considered the time from model prediction (when the risk score crosses the prediction threshold) to sepsis for the ED and ICU patient populations, since it’s possible to have a sepsis prediction model with high performance but with no lead time (for instance by incorporating treatment information as a feature in the model).

Given that the COMPOSER algorithm was designed to be generalizable, we postulated that sensitivity and specificity of the COMPOSER score would not vary significantly, assuming that the physiological characteristics of septic compared to non-septic patients would remain similar, across the various cohorts. Based on preliminary observations from our development dataset we anticipated that at 80% sensitivity COMPOSER would achieve at least 90% specificity across various cohorts. Additionally, we observed a sepsis (hourly window-wise) incidence rate of roughly 3.0–4.0% in the ICU and 2.0–5.5% in the ED. Plugging these incident rates into the standard equations for PPV (and NPV) as a function of sensitivity and specificity yields approximate values in the range of 20.0–25.0% (and NPV of 99.0–99.3%) in the ICU and 14.0– 30.0% (and NPV of 98.7-99.5%) in the ED, respectively. As such, we formed two hypotheses for the ICU and ED populations as follows. We hypothesized that across the various ICU patient cohorts, with 95% power (beta=0.05; type-II error) at an alpha of 5% (type-I error) with sufficient sample size^34^, we can show that the COMPOSER score achieves at least 20% PPV and 98% NPV. Similarly, we hypothesized that across the various ED patient cohorts, with 95% power (beta=0.05; type-II error) at an alpha of 5% (type-I error) with sufficient sample size we can show that the COMPOSER score achieves at least 10% PPV and 98% NPV. (Please see Appendix F of Supplementary Material for sample size calculations). Additionally, we hypothesized that COMPOSER yields higher AUC, PPV, NPV and DOR compared to competing models (GB-Vital, baseline FFNN and ESPM models) as described next. All baseline models (including GB-Vital) were trained using the same sepsis criteria for consistency. The only exception was the ESPM risk model, since we only had access to the ESPM risk scores in our electronic health record (not the actual model).

#### GB-Vital

This is a replication of the sepsis detection model as proposed by Mao et al.^6,15^.The model corresponds to a gradient-boosted classifier of decision trees built using six vital signs measurements: systolic blood pressure, diastolic blood pressure, heart rate, respiratory rate, oxygen saturation and temperature.

#### FFNN

This model corresponds to a 2 layer feedforward neural network (of size 40 and 25) that uses the same number of input features as that of COMPOSER.

#### ESPM

This model corresponds to the Epic’s commercially available Best Practice Advisory (BPA) alert^35^. We only had access to the risk scores produced by this system at Hospital-A during the temporal validation time-frame.

### Data processing, training and hyperparameters

First, the combined Hospital-A ICU and ED training set was standardized by first applying normalization transformations, followed by subtracting the mean and dividing by the standard deviation. Next, all remaining datasets were normalized using exactly the same transformations utilized in the training data. For handling missing data, we used a simple sample-and-hold approach in all the datasets, with mean-imputation at the start of all time series records.

#### Weighted input layer

The scaling factors *α*_*i*_ were all initialized to 1. *Model:* The learning rates for encoder, sepsis predictor and domain classifier were set to 0.01. To minimize overfitting and to improve generalizability of the model, L1–L2 regularization was used with L2 regularization parameter set to 1e-3 for encoder and sepsis predictor, 1e-4 for domain classifier and L1 regularization parameter set to 1e-3 for encoder and sepsis predictor, 1e-4 for domain classifier.

Mini-batch size for the source dataset was fixed at a total of 10000 windows (50% septic windows, 50% non-septic windows). Mini-batch size for the target dataset was set at 5000 windows. The encoder, sepsis predictor, and domain classifiers were each composed of a single layer neural network of dimensions 40, 25, and 25 neurons, respectively. Both the sepsis predictor and domain classifier were further followed by a fully connected layer and a softmax layer.

#### Conformal predictor

Threshold ∈ was set at 0.05. Trust set consisted of an equal proportion of septic windows and non-septic windows, optimized to minimize the deleterious effects of examples with large numbers of missingness on prediction performance (see Appendix E.3 of Supplementary Material). COMPOSER was trained for a total of 500 epochs using Adam optimizer^36^, with early stopping. All hyper-parameters of the model (number and size of layers for encoder-sepsis predictor-domain classifier, learning rate, mini-batch size, L1 regularization parameter, and L2 regularization parameter) were optimized using Bayesian optimization on the validation set of the development site^37^. All pre-processing of data was performed using Numpy^38^ with the rest of the pipeline implemented using TensorFlow^39^.

